# Hyperacute effects of non-code dose bolus epinephrine in pediatric cardiac intensive care patients: insights from high fidelity physiologic data

**DOI:** 10.1101/2025.07.01.25330684

**Authors:** Wesam Sourour, Michael Evans, Kevin Le, Saul Flores, Juan S. Farias, Rohit S. Loomba

**Affiliations:** Ann & Robert H. Lurie Children’s Hospital, Chicago, IL, USA; Northwestern University Feinberg School of Medicine, Chicago, IL, USA; Texas Children’s Hospital, Baylor College of Medicine, Houston, TX, USA; Children’s Mercy Hospital, Kansas City, MO, USA

## Abstract

**Background:** Non-code dose boluses of epinephrine are utilized in critically ill pediatric patients during periods of hemodynamic deterioration, often with the hopes of preventing a cardiac arrest. Data regarding the physiologic effects of these administrations are limited. The primary aim of this study was to use high fidelity physiologic data to characterize the effects of intravenous non-code dose bolus epinephrine.

**Methods:** Pediatric patients in the cardiac intensive care unit who received non-code dose bolus epinephrine were identified. Those who received fluid boluses or chest compressions within 2 minutes of bolus epinephrine were excluded. ARIMAX analyses were conducted to characterize the time-dependent changes in hemodynamic indices. Cluster analyses were then conducted to determine patterns in hemodynamic changes associated with bolus epinephrine.

**Results:** A total of 71 non-code dose bolus epinephrine administrations were included in the final analyses. Heart rate, blood pressure, and renal near infrared spectroscopy all demonstrated statistically significant changes after bolus epinephrine administration. Peak change in each was 40%, 52%, and 9%, respectively, with peaks occurring between 60-seconds and 120-seconds after administration. Three response-based clusters were identified.

**Conclusion:** Non-code dose bolus epinephrine is associated with a significant increase in heart rate, blood pressure, and systemic oxygen delivery. Cluster analysis using the peak change identified distinct clinical clusters.

## Introduction

Epinephrine is a frequently used vasoactive in the pediatric cardiac intensive care unit ^1,2^. It is often utilized as a continuous infusion but can also be used as a bolus dose. Bolus doses of epinephrine were initially utilized during cardiopulmonary resuscitation, although boluses have also started being used in deteriorating patients who are not receiving cardiopulmonary resuscitation. In this setting, the clinical aim is often to avoid cardiorespiratory arrest ^3,4^.

When epinephrine is used in the non-arrest setting, it is often used at a lower dose. There are limited data of this non-code dose bolus epinephrine, although its use is largely endorsed by pediatric intensivists ^5^. Its use has even been mentioned in the scientific statement regarding cardiopulmonary resuscitation in infants and children with cardiac disease ^6^. The limited data suggest that non-code dose bolus epinephrine may significantly raise blood pressure and heart rate in children.

The primary aim of this study was to characterize the change in heart rate, blood pressure, and renal near infrared spectroscopy in the 2 minutes following administration of non-code dose bolus epinephrine in pediatric cardiac intensive care patients.

## Methods

### Study design

This was a single center, retrospective study using high fidelity physiologic data collected with the Sickbay platform (Medical Informatics Company, Houston, TX, USA). The primary aim of the study was to quantify the response in heart rate, systolic arterial blood pressure, mean arterial blood, diastolic arterial blood pressure, central venous pressure, near infrared spectroscopy, and arterial saturation in the 120 seconds (2 minutes) following administration of non-code dose bolus epinephrine in non-cardiorespiratory arrest situations in the cardiac intensive care unit. Secondary aims of the study were to quantify the magnitude and timing of peak effect of non-code dose bolus epinephrine on these physiologic parameters and determine if age, weight, or circulatory pattern influenced the magnitude of response.

Institutional review board approval was obtained for this study.

### Variables of interest

The following physiologic data were collected from the Sickbay platform with 1-second temporal resolution: heart rate (beats per minute), systolic arterial blood pressure (mmHg), mean arterial blood pressure (mmHg), diastolic arterial blood pressure (mmHg), central venous pressure (mmHg), renal near infrared spectroscopy (%), and arterial saturation (%). The data for heart rate was sourced from telemetry, data for arterial blood pressure from an arterial line, data for central venous pressure from a central venous line, and renal near infrared spectroscopy from the INVOS 5100 (Medtronic, Boston, MA, USA).

Clinical variables collected from the electric medical record included age (months), weight (kg), and circulatory type at time of non-code dose bolus epinephrine administration.

Principle circulatory types were as follows: fully septated biventricular, left to right shunt, limitation to pulmonary blood flow, limitation to systemic blood flow, preoperative transposition, functionally univentricular (multidistributive, Glenn, or Fontan), or ventricular dysfunction. It is possible that some patients were able to be classified in more than one circulatory type but the principle one was selected based on clinical judgement.

### Administration inclusion

Non-code dose bolus epinephrine administrations in the pediatric cardiac intensive care unit from January 1, 2022 to May 1, 2025 were identified. Administrations to patients who did not receive cardiopulmonary resuscitation, code-dose vasoactive administration, sodium bicarbonate, or endotracheal intubation in the two minutes prior and after were eligible for inclusion. Administrations to patients that received any of these interventions in the period of interest, did not have Sickbay data available, or did not have an arterial line data during the administration were excluded.

### Statistical analyses

Continuous variables are reported as mean and standard deviation while categorical variables are reported as absolute count and percent.

Missing data were identified, and median imputation was performed to handle missing values.

Next, the timing of non-code dose bolus epinephrine administration was annotated into the data for each administration. The timestamps in the data were used to determine relative times in seconds for all data points per administration. Thus, relative time was negative for timepoints prior to non-code dose bolus epinephrine administration and positive for timepoints after.

For each administration the maximal percent peak change in heart rate, mean arterial blood pressure, and renal near infrared spectroscopy was determined from the time series. The relative time at which this occurred was also determined.

Next, autoregressive integrated moving average with exogenous inputs analyses were performed. Time-series modeling was conducted using autoregressive models to quantify the effect of non-code-dose bolus epinephrine administration on physiologic parameters while adjusting for age, weight, and circulatory type. Separate models were fit for heart rate, systolic arterial blood pressure, mean arterial blood pressure, diastolic blood pressure, central venous pressure, renal near infrared spectroscopy, arterial saturation and respiratory rate. For each model, the dependent variable was the binned physiologic time series, and the independent variables were phase (before or after epinephrine), age, weight, and circulatory type. The models used an autoregressive lag of 1 second and a moving average lag of 1 second. Parameters were estimated using maximum likelihood using the SARIMAX implementation in the statsmodels Python library. Model fit was assessed.

Next, unsupervised cluster analysis were performed to identify distinct patterns of physiologic response to non-code dose bolus epinephrine. An unsupervised clustering approach was performed on the time series of percent change in heart rate, mean arterial blood pressure, and renal near infrared spectroscopy from baseline. A dynamic time warping distance matrix was computed across administrations to account for potential phase shifts in the timing of peak responses. Clustering was then performed using agglomerative hierarchical clustering with Ward’s linkage. The optimal number of clusters was determined using silhouette scores. Descriptive characteristics including age, weight, and circulatory type were compared across clusters using analysis of variance or Kruskal-Wallis tests, as appropriate.

Finally, a regression analysis was conducted to model percent change in mean arterial blood pressure and percent change in renal near infrared spectroscopy separately in only those with functionally univentricular multidistributive circulation.

All statistical analyses were conducted using a local Python environment. A p-value of less than 0.05 was considered statistically significant. All use of the word “significant” or “significantly” in this manuscript refers to statistical significance unless explicitly specified to be clinical significance.

## Results

### Cohort characteristics

A total of 71 non-code dose bolus administrations were included in the final analyses. The average age at non-code dose bolus epinephrine administration was 36.5 months and the average weight was 13.7kg. The most frequent circulatory type was functionally univentricular multidistributive circulation in 28 (39%) patients followed by fully septated biventricular circulation in 22 (31%) patients (Table 1). The dose of epinephrine was 1 mcg/kg for all patients, as this dose is standardized in the cardiac intensive care unit.

**Table 1.** cohort characteristics, administration level data.

|  |  |
| --- | --- |
| Age (months) | 36.5 ± 69.8 |
| Weight (kg) | 13.7 ± 19.5 |
| Circulatory type |  |
| Fully septated biventricular | 22 (31%) |
| Left to right shunt | 3 (4%) |
| Limitation to pulmonary blood flow | 4 (6%) |
| Limitation to systemic blood flow | 2 (3%) |
| Preoperative transposition | 4 (6%) |
| Functionally univentricular multidistributive | 28 (39%) |
| Ventricular dysfunction | 8 (11%) |

### Physiologic effects

Figures 1 through 8 characterize the percent change in the physiologic variables of interest with relative time. The moment of non-code dose bolus epinephrine administration is time 0. Peak change and time of peak change is characterized for each physiologic variable in table 2. The greatest percent change was noted in central venous pressure which demonstrated a peak change of 187.3% at 120 seconds after epinephrine administration. Systolic arterial blood pressure demonstrated the second greatest peak change at 57.8% at 53 seconds. Renal near infrared spectroscopy demonstrated a peak change of 9.4% at 72 seconds.

**Table 2.** Percent peak change and time at peak change for each physiologic variable after bolus epinephrine administration.

|  | **Peak change (%)** | **Time to peak (s)** |
| --- | --- | --- |
| ***Heart rate*** | 40.5 | 120 |
| ***Systolic arterial blood pressure*** | 57.8 | 53 |
| ***Mean arterial blood pressure*** | 52.5 | 60 |
| ***Diastolic arterial blood pressure*** | 56.6 | 115 |
| ***Central venous pressure*** | 187.3 | 120 |
| ***Renal near infrared spectroscopy*** | 9.4 | 72 |
| ***Arterial saturation*** | 17.1 | 116 |
| ***Respiratory rate*** | -3.4 | 116 |

Autoregressive mean integrated average analyses demonstrated that a significant percent increase in all physiologic variables after non-code dose bolus epinephrine except respiratory rate which demonstrated no significant change (table 3). Effect of age, weight, and circulatory type on the percent change of physiologic variables is also outlined in table 3. Of note is that functionally univentricular multidistributive circulation tended to be associated with less percent increase in arterial blood pressure, less percent increase in renal near infrared spectroscopy, and less percent increase in respiratory rate.

**Table 3.** Results from the autoregressive integrated moving average analyses characterizing only significant associations between change in the physiologic variables of interest and the independent variables.

| **d** | **Bolus epinephrine** | **Age** | **Weight** | **Circulatory type** |
| --- | --- | --- | --- | --- |
| ***Heart rate*** | Increased | Decreased | Decreased | No significant association |
| ***Systolic arterial blood pressure*** | Increased | Increased | Increased | Decreased with functionally univentricular multidistributive |
| ***Mean arterial blood pressure*** | Increased | Increased | Increased | Decreased with functionally univentricular multidistributive |
| ***Diastolic arterial blood pressure*** | Increased | Increased | Increased | Decreased with functionally univentricular multidistributive |
| ***Central venous pressure*** | Increased | No significant association | No significant association | No significant association |
| ***Renal near infrared spectroscopy*** | Increased | No significant association | No significant association | Increased with functionally univentricular multidistributive |
| ***Arterial saturation*** | Increased | No significant association | No significant association | Increased with functionally univentricular multidistributive |
| ***Respiratory rate*** | No significant association | Decreased | Decreased | Increased with functionally univentricular multidistributive |

**Table 4.** percent change in physiologic variable across clusters.

|  | **Cluster 1** | **Cluster 2** | **Cluster 3** |
| --- | --- | --- | --- |
| ***Heart rate*** | 9.6 ± 8.3 | 30.0± 22.0 | 15.9 ± 43.4 |
| ***Mean arterial blood pressure*** | 45.3 ± 27.9 | 63.4 ± 28.5 | 116.5 ± 72.3 |
| ***Near infrared spectroscopy*** | 4.3 ± 5.5 | 4.0 ± 3.5 | 26.3 ± 11.8 |

### Cluster analyses

Unsupervised cluster analyses using percent change in heart rate, mean arterial blood pressure, renal near infrared spectroscopy, weight, age, and circulatory type identified three distinct clusters. Model quality was 0.56, identifying strong cluster segregation. The percent change in mean arterial blood pressure had greatest feature importance at 0.29 followed by percent change in renal near infrared spectroscopy with a feature importance of 0.24, and then percent change in heart rate with a feature importance of 0.21. Weight, age, and circulatory type all had a feature importance of less than 0.1.

Cluster 1 had a mean age of 11 months and consisted of many patients that had fully septated, biventricular hearts but also contained some with functionally univentricular multidistributive circulation. Cluster 1 patients demonstrated the lowest percent increase in heart rate and mean arterial blood pressure.

Cluster 2 had a mean age of 147 months and consisted of many patients with ventricular dysfunction. Cluster 2 demonstrated the greatest percent increase in heart rate, second greatest percent increase in mean arterial blood pressure, and the lowest increase in the renal near infrared spectroscopy.

Cluster 3 had a mean age of 3.5 months and consisted of many patients with functionally univentricular multidistributive circulation and preoperative transposition. Cluster 3 demonstrated the second greatest percent increase in heart rate, greatest increase in mean arterial blood pressure, and greatest increase in renal near infrared spectroscopy.

### Regression analysis for patients with functionally univentricular multidistributive circulation

Regression analyses were conducted to model peak change in systolic blood pressure and renal near infrared spectroscopy. The regression for peak change in systolic blood pressure demonstrated that the only predictor was baseline arterial blood pressure with a lower baseline blood pressure being associated with a greater peak increase (beta-coefficient - 2.18, p= 0.04). The regression for peak change in renal near infrared spectroscopy demonstrated that the only predictor was weight with higher weight being associated with lower peak change in renal near infrared spectroscopy (beta-coefficient -3.37, p= 0.04).

## Discussion

This study used high-fidelity physiologic data to characterize the effects of non-code dose bolus epinephrine for the first two minutes after administration. To highlight key findings, mean arterial blood pressure increased 52%, heart rate increased 40%, and renal near infrared spectroscopy increased 9% in all patients. Most of these effects peaked in the observed time period. These findings were mediated by epinephrine as demonstrated by ARIMA analyses accounting for age, weight, and circulatory type.

Findings in patients with functionally univentricular multidistributive circulations were particularly interesting. The ARIMA demonstrated that those with functionally univentricular hearts tended to have a decreased increase in blood pressure, increased change in renal near infrared spectroscopy, and increased change in arterial saturation. Cluster analyses demonstrated a distinct cluster of primarily functionally univentricular multidistributive circulation patients (although it did not contain all these patients). The children in this cluster had greater increases in heart rate, blood pressure, and near infrared spectroscopy, which means there were some apparent discrepancies in findings between the ARIMAX and cluster analyses. Because of this disagreement, an additional regression analysis was conducted for only patients with functionally univentricular multidistributive circulation. To reconcile these findings, it is important to highlight that the ARIMAX demonstrates a blunted response in arterial blood pressure in those with functionally univentricular multidistributive circulation, but the clusters demonstrate that while this is a trend that there are some patients who have a greater response. The subsequent regression of just these particular patients helped identify what patients did demonstrate a greater response. Given these data, these different analyses are all complementary.

The findings of this study indicate that non-code-dose bolus epinephrine is associated with improved hemodynamic state. The increase in near infrared spectroscopy indicates improvement in systemic oxygen delivery as observed by an increase in the renal near infrared spectroscopy values^7–9^. This improvement is likely due to an increase in cardiac output which then yields an increase in oxygen content and delivery in pediatric patients ^10,11^. The increase in blood pressure is also, in part, mediated by such an increase in cardiac output although the degree to which it is augmented by flow and resistance is not easily discernible by these data. The mechanism of action of epinephrine is well demonstrated and consists of both alpha- and beta-receptor agonism. The relative proportion of alpha- and beta-agonism appears to be dose-dependent. At the 1 mcg/kg dosing utilized in the current study, epinephrine should have both alpha- and beta-agonism with beta-agonism expected to be slightly more predominant. Beta-1 agonism at this dose would lead to increased heart rate, contractility, and atrioventricular nodal conduction, beta-2 agonism at this dose would lead to skeletal muscle vasodilation and some bronchodilation ^12^.

The magnitude of change in heart rate and blood pressure noted in this study is similar to findings from previous investigations. Two previous studies used a 5-minute window before and after to observe for effects. Given the sheer amount of data collected for each event, along with the enhanced temporal resolution of the data the quick effect of non-code dose bolus epinephrine, and the clinical aims for the use of non-code dose bolus epinephrine, it was felt that a 2-minute before-and-after window was reasonable in this study. In comparison to this study, Ross and colleagues demonstrated a 22% increase in mean arterial blood pressure and 5% increase in heart rate at the 5-minute mark ^13,14^. A separate study by Reiter and colleagues with variable dosing demonstrated that there was markedly lower magnitude of effect with epinephrine doses of under 1 mcg/kg ^15^.

Use of non-code dose bolus epinephrine has been endorsed by many pediatric intensivists with an overwhelming majority saying they already use or would consider using it. Ross and colleagues conducted a survey of 63 intensivists across 35 institutions with 94% of those surveyed responding that they would consider using non-code dose bolus epinephrine in deteriorating patients not requiring cardiopulmonary resuscitation. The 6% of those who responded to the survey saying they would not use non-code dose bolus epinephrine cited a lack of evidence or unfamiliarity as their reasons for not considering its use. Amongst those surveyed that were already utilizing non-code dose epinephrine, there was a reported large variation in dosing and vernacular. The reported doses were quite variable, ranging from 0.1 mcg/kg to 10 mcg/kg with the most common dose being 1 mcg/kg among 68% of those who responded ^5^. The general endorsement and adoption of non-code dose bolus epinephrine is reflected as a recommendation in the scientific statement regarding cardiopulmonary resuscitation in infants and children with cardiac disease, stating that is reasonable to utilize such doses in the peri-arrest patient ^6^.

The present study adds to the limited data available regarding the effects of non-code dose bolus epinephrine in pediatric patients. The data are sourced in high fidelity, allowing for highly powered statistical analyses able to detect even small but significant effects. The exclusion criteria utilized also provide a setting in which the effects can be more or less isolated to be the result of the non-code dose bolus epinephrine. While this study has its strengths, it also has its limitations. This is a single-center study. Due to local practice, we were unable to identify the effect of dose, as a uniform dose is universally utilized, but the uniformity of dosing also serves as a strength. Additionally, this study only looked at the effect in those who did not experience a cardiac arrest within 2 minutes of the administered dose. It is plausible that those who do arrest in this time period may have a different response to non-code dose bolus epinephrine. Despite its limitations, this study contains provides novel insight into the effects of non-code dose bolus epinephrine, especially illustrating differences in response in patients with functionally univentricular multidistributive circulation.

## Conclusion

Non-code dose bolus epinephrine is associated with a significant increase in heart rate, blood pressure, and systemic oxygen delivery. Peak change in each was 40%, 52%, and 9%, respectively with peaks occurring between 60-seconds and 120-seconds after administration. Cluster analysis using the peak change identified distinct clinical clusters.

## Data Availability

Data are not publicly available

---

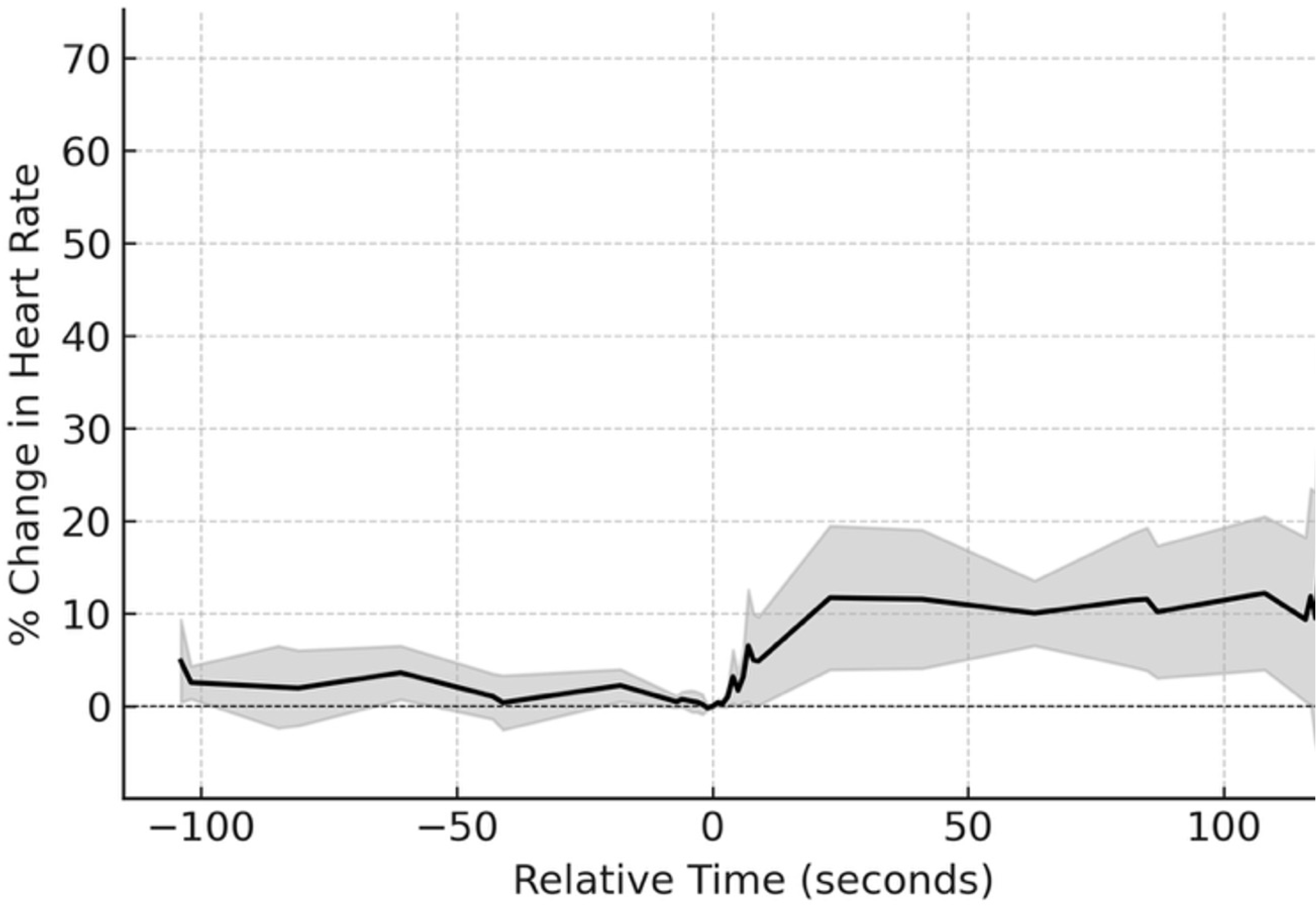
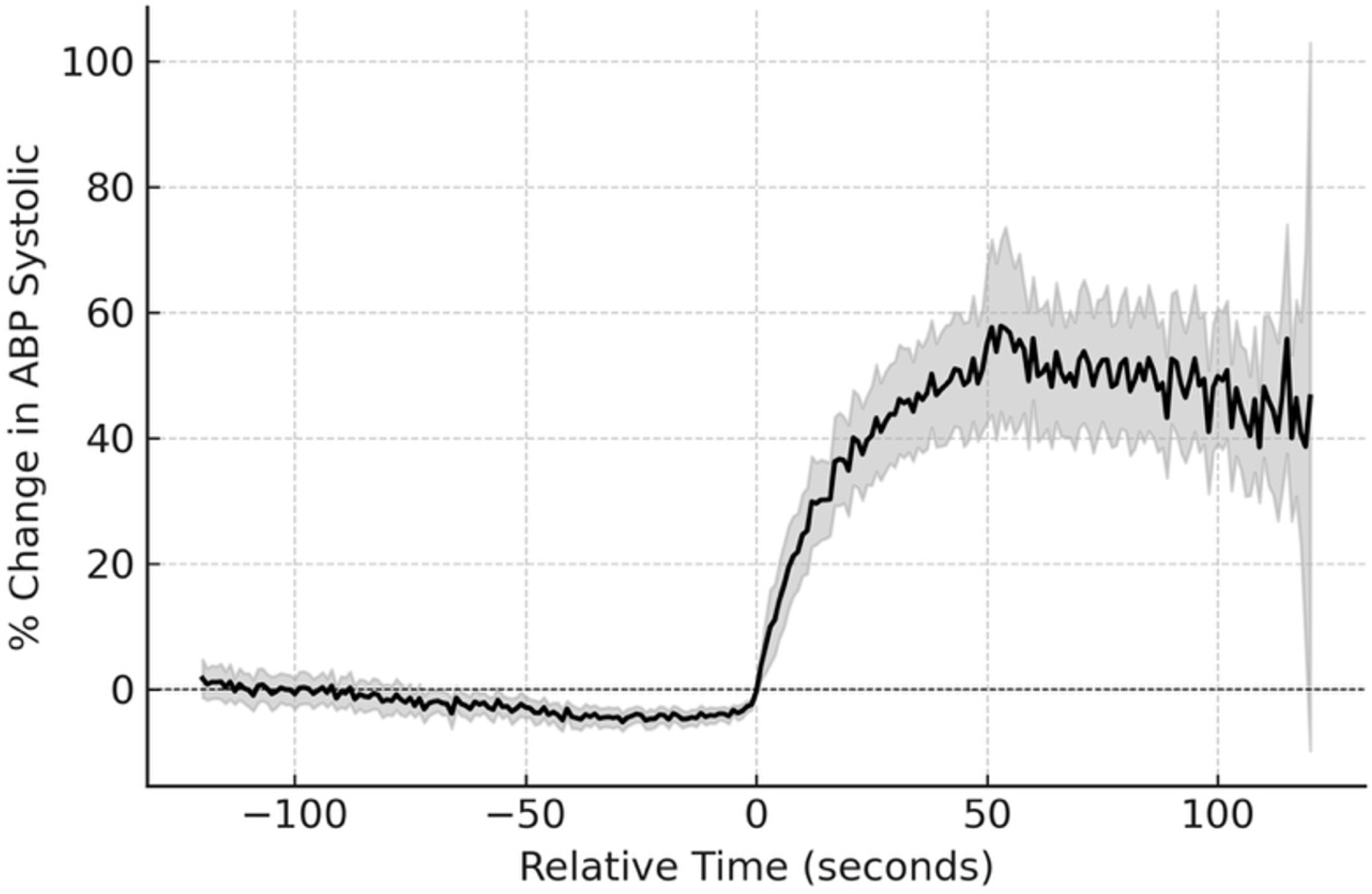
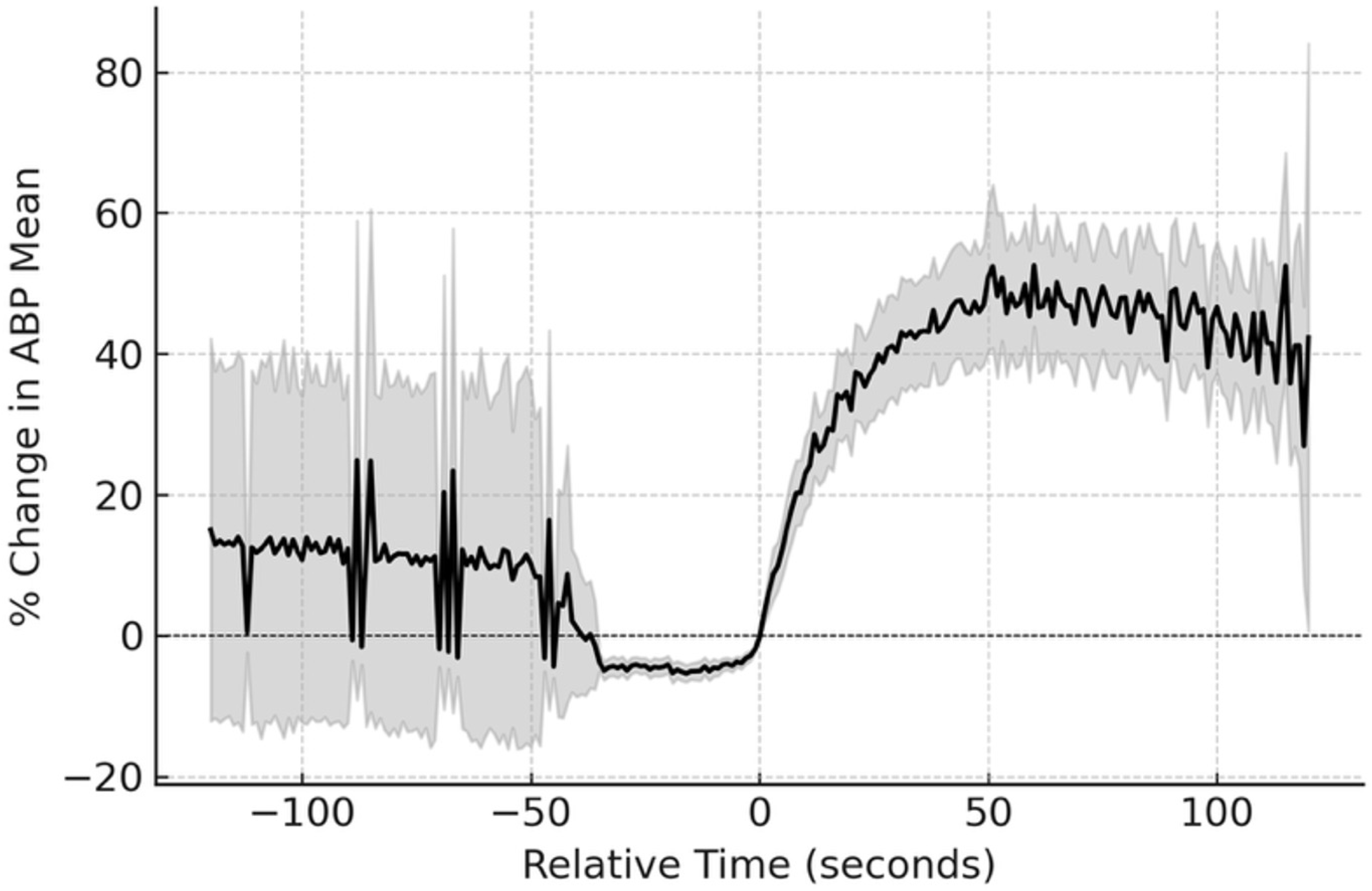
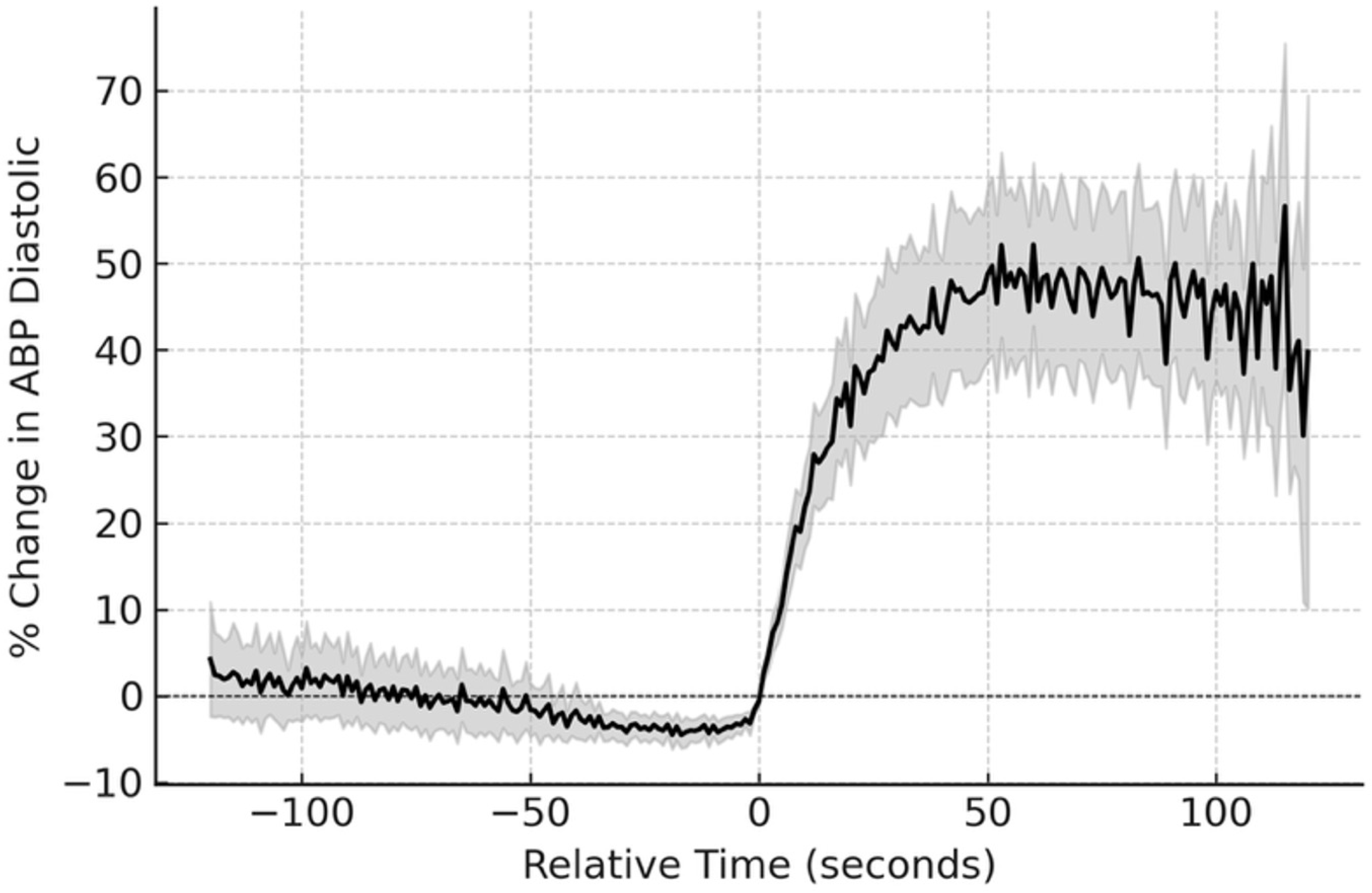
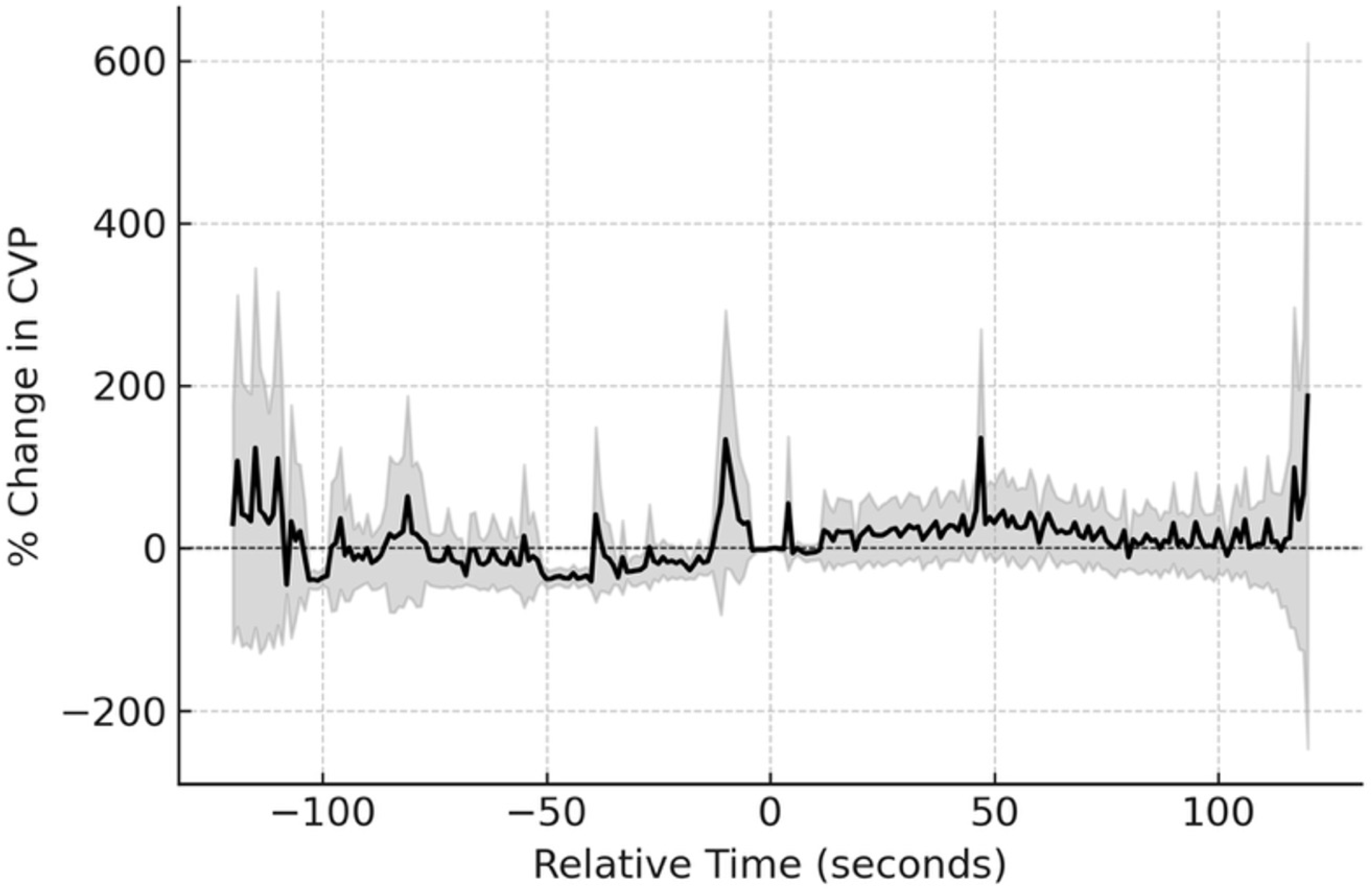
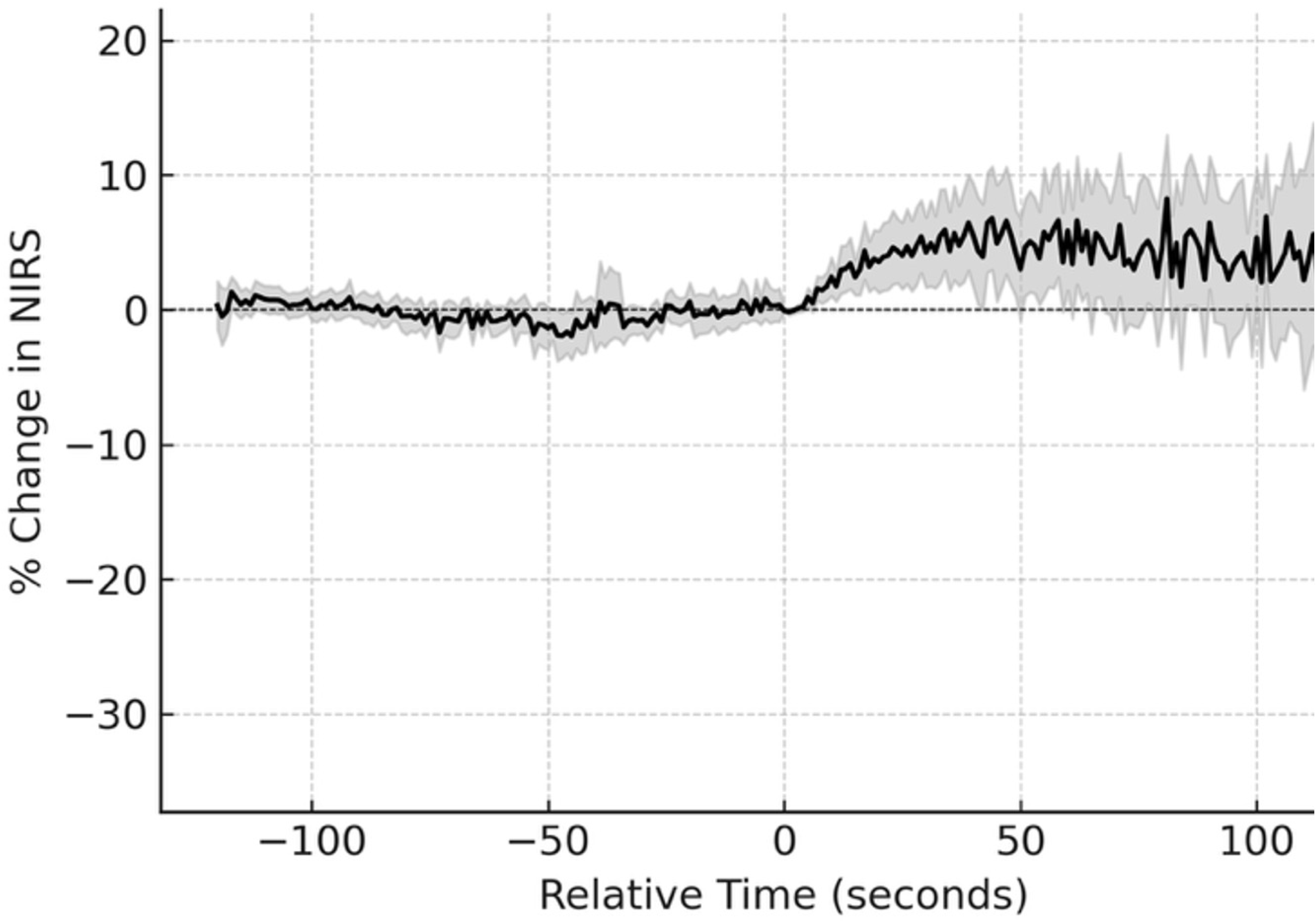
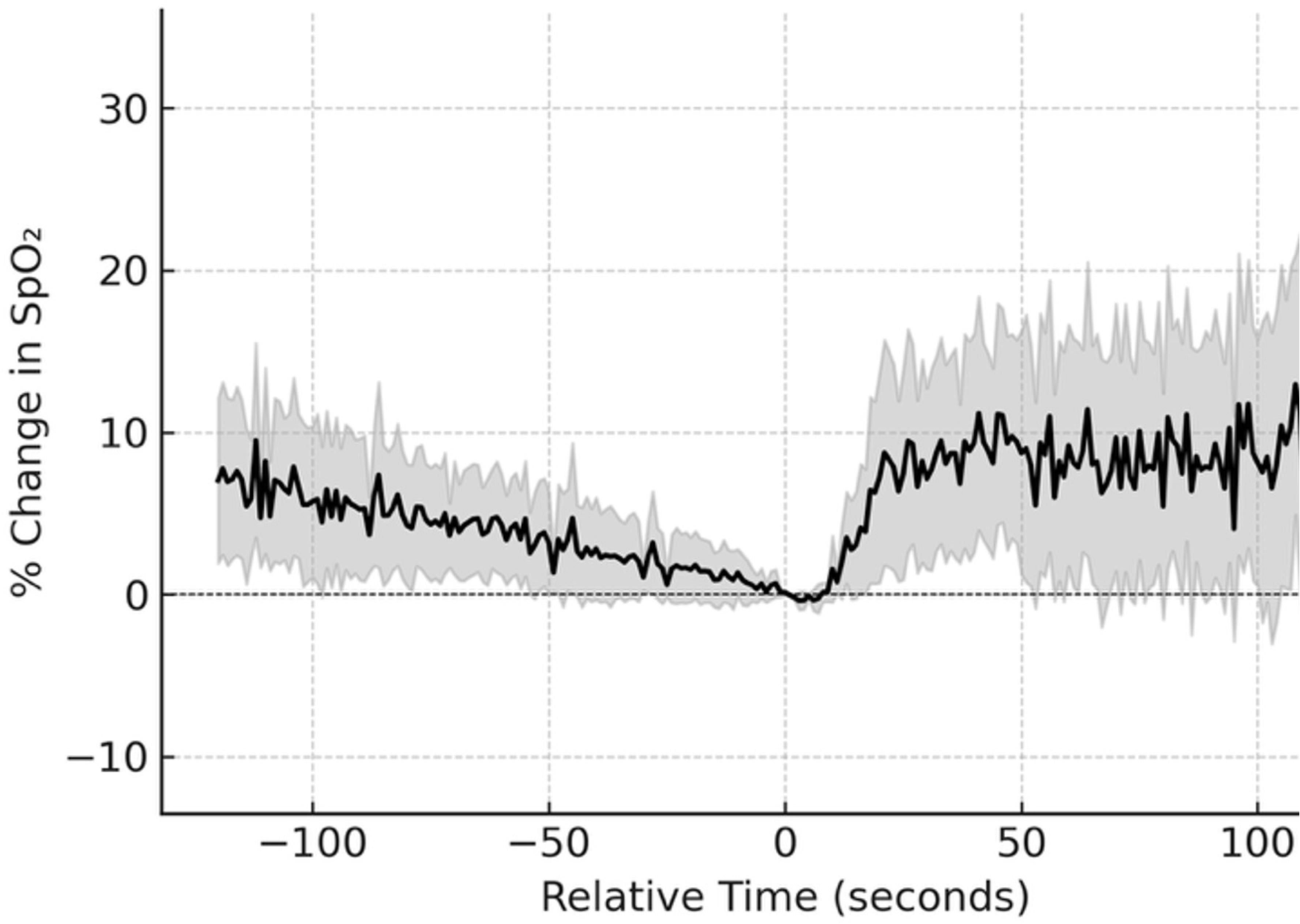
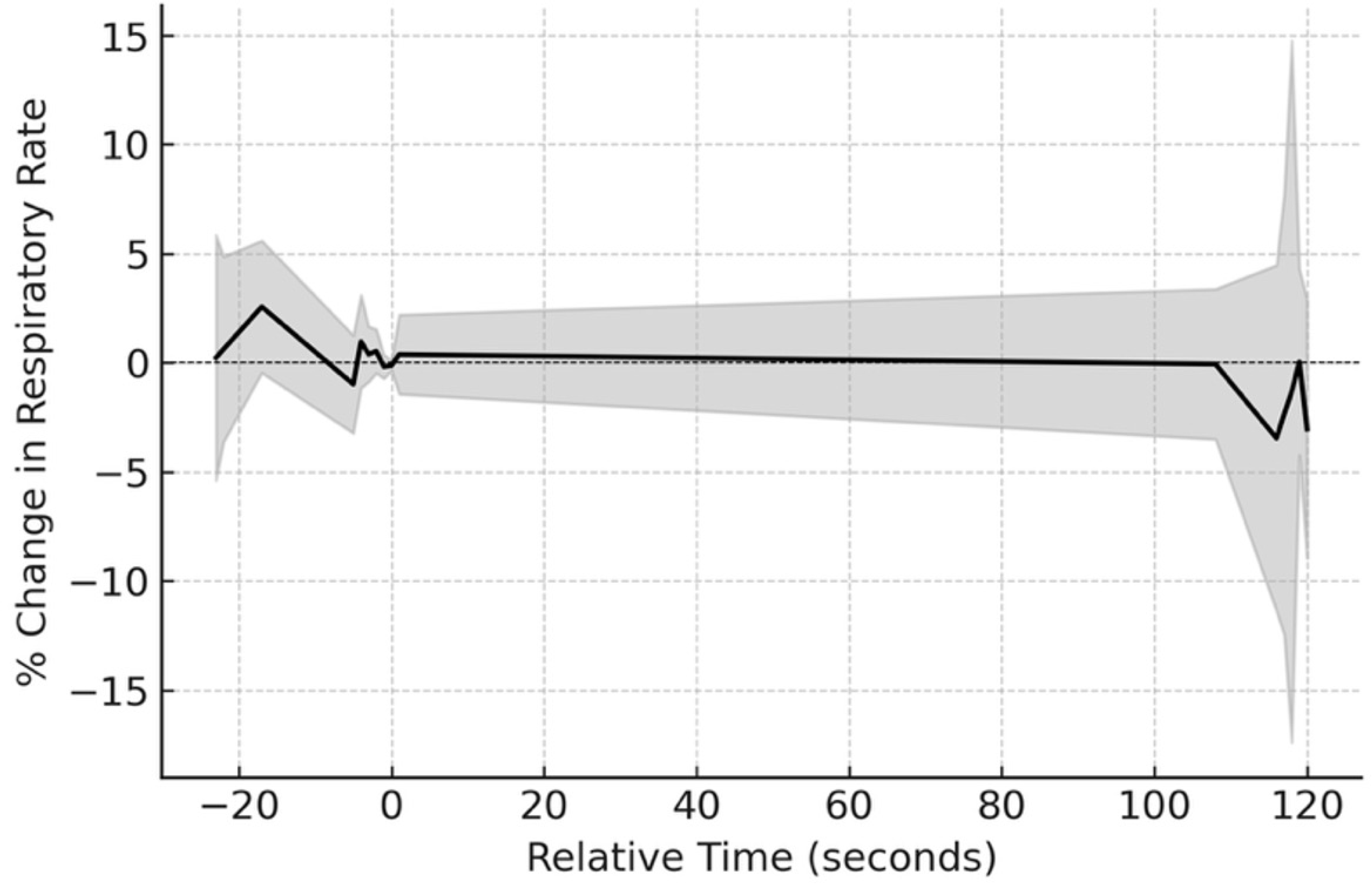

